# Fasting alters the gut microbiome with sustained blood pressure and body weight reduction in metabolic syndrome patients

**DOI:** 10.1101/2020.02.23.20027029

**Authors:** András Balogh, Hendrik Bartolomaeus, Ulrike Löber, Ellen G. Avery, Nico Steckhan, Lajos Markó, Nicola Wilck, Ibrahim Hamad, Urša Šušnjar, Anja Mähler, Christoph Hohmann, Holger Cramer, Gustav Dobos, Till Robin Lesker, Till Strowig, Ralf Dechend, Danilo Bzdok, Markus Kleinewietfeld, Andreas Michalsen, Dominik N. Müller, Sofia K. Forslund

**Affiliations:** Experimental and Clinical Research Center (ECRC), a cooperation of Charité - Universitätsmedizin Berlin and Max Delbruck Center for Molecular Medicine (MDC), 13125 Berlin, Germany; Charité-Universitätsmedizin Berlin, corporate member of Freie Universität Berlin, Humboldt-Universität zu Berlin, and Berlin Institute of Health, 10117 Berlin, Germany; Berlin Institute of Health (BIH), 10178 Berlin, Germany; DZHK (German Centre for Cardiovascular Research), partner site Berlin, 13316 Berlin, Germany; Max Delbruck Center for Molecular Medicine in the Helmholtz Association (MDC), 13125 Berlin, Germany; Department of Biology, Chemistry, and Pharmacy, Freie Universität Berlin, 14195 Berlin, Germany; Department of Internal and Integrative Medicine, Immanuel Krankenhaus Berlin, 14109 Berlin, Germany; VIB Laboratory of Translational Immunomodulation, VIB Center for Inflammation Research (IRC), UHasselt, Campus Diepenbeek, 3590 Hasselt, Belgium; Department of Internal and Integrative Medicine, Kliniken Essen-Mitte, Faculty of Medicine, University of Duisburg-Essen, 45136 Essen, Germany; Department of Microbial Immune Regulation, Helmholtz Centre for Infection Research, 38124 Braunschweig, Germany; Hannover Medical School, 30625 Hannover, Germany; Department of Cardiology and Nephrology, HELIOS-Klinikum, 13125 Berlin, Germany; Department of Psychiatry, Psychotherapy, and Psychosomatics, Rheinisch-Westfälische Technische Hochschule (RWTH) Aachen University, 52072 Aachen, Germany; Jülich Aachen Research Alliance (JARA), Translational Brain Medicine, 52056 Aachen, Germany; Parietal Team, Institut National de Recherche en Informatique et en Automatique (INRIA), Neurospin, Commissariat à l’Energie Atomique (CEA) Saclay, 91191 Gif-sur-Yvette, France

**Keywords:** metabolic syndrome, hypertension, periodic fasting, microbiome, immunome

## Abstract

Periods of fasting and refeeding may reduce cardiometabolic risk elevated by Western diet. We show that in hypertensive metabolic syndrome (MetS) patients (n=35), a 5-day fast followed by a modified DASH diet (Dietary Approach to Stop Hypertension) reduced systolic blood pressure (SBP), antihypertensive medication need, and body-mass index (BMI) at three months post intervention compared to a modified DASH diet alone (n=36). Fasting altered the gut microbiome, impacting bacterial taxa and gene modules associated with short-chain fatty acid production. Cross-system analyses revealed a positive correlation of circulating mucosa-associated invariant T (MAIT) cells, non-classical monocytes and CD4+ effector T cells with SBP. Furthermore, regulatory T cells (Tregs) positively correlated with BMI and weight. Machine learning could predict sustained SBP-responsiveness within the fasting group from baseline immunome data, identifying CD8+ effector T cells, Th17 cells and Tregs as important contributors to the model. The high-resolution multi-omics data highlights fasting as a promising non-pharmacological intervention in MetS.

---

Fasting can prolong survival and reduce disease burden in rodent models, and possibly in humans^1^. In contrast, today’s Western diet promotes cardiometabolic disease^2^. The relationship how diet affects the gut microbiota, immune system and subsequently host (patho)physiology is not fully understood, and information is lacking on how periodic fasting affects the gut microbiome in patients with metabolic syndrome (MetS). To reduce cardiometabolic disease (CMD) risk, exercise and a healthy diet are often prescribed. Shifting from a ‘Western diet’ to a healthier ‘Mediterranean-like’ DASH diet^3^ to achieve optimal nutrition and negative energy balance is recommended, although compliance is a major hurdle. Our study is the first of its kind to investigate the effects of a lifestyle modification in combination with fasting therapy in patients with MetS using a multi-omics approach. The ‘Western diet’ is known to induce metabolic inflammation, accelerating CMD^4^. The gut microbiota is a delicate ecosystem that plays a pivotal role in health and disease. Dysbiosis has been observed as a characteristic of several inflammatory, cardiovascular, and metabolic disorders (e.g. obesity)^5^, including hypertension^6, 7^. The ‘healthy’ gut microbiome is relatively stable, although various factors such as antibiotics, intestinal infections, profound dietary or lifestyle changes, such as moving on or off a ‘Western diet’ can induce transient or persistent changes to this ecosystem. Traditionally, fasting plays an important role in different cultural and religious practices. Dramatic caloric restriction not only affects host health and physiology, but likely also has an impact on the microbiome. However, the effect of fasting on the gut microbiome and the consequences of refeeding after such an intervention have not yet been described.

## Methods

### Study design and patients

Subjects diagnosed with MetS and elevated systolic blood pressure (BP) were randomly allocated into two intervention groups. The fasting arm underwent five days of fasting according to ‘Buchinger’s protocol’^8^ followed by three months of a modified DASH diet (also referred to as the refeeding period) (*Figure 1A, Table 1*). The other arm underwent a modified DASH intervention only (referred to as DASH). Ethical approval was obtained from the local ethics committee (No. EA4/141/13). Informed consent was obtained from all subjects before entering the study. The present study complies with the Declaration of Helsinki and was registered at ClinicalTrials.gov (NCT02099968). For further methodological and statistical details, see Supplementary information online.

**Table 1.** Patient characteristics at baseline.

|  | **FASTING**<br>**+ DASH** | **DASH** |
| --- | --- | --- |
| Females/Males | 23/12 | 21/15 |
| Age (year) | 58 ± 8 | 62 ± 8 |
| Height (cm) | 171 ± 8 | 171 ± 9 |
| Office SBP (mm Hg) | 136 ± 15 | 138 ± 16 |
| Office DBP (mm Hg) | 88 ± 11 | 88 ± 9 |
| 24h ABPM SBP (mm Hg) | 132 ± 9 | 131 ± 9 |
| 24h ABPM DBP (mm Hg) | 81 ± 8 | 81.4 ± 7 |
| 24h ABPM MAD (mm Hg) | 104 ± 8 | 104 ± 7 |
| 24h ABPM peripheral resistance (mm Hg*s/ml) | 1.4 ± 0.1 | 1.3 ± 0.1 |
| SBP day (mm Hg) | 134 ± 10 | 133 ± 10 |
| DBP day (mm Hg) | 83 ± 9 | 84 ± 7 |
| SBP nocturnal (mm Hg) | 120 ± 12 | 121 ± 10 |
| DBP nocturnal (mm Hg) | 71.5 ± 8 | 71.6 ± 7 |
| Weight (kg) | 99 ± 17 | 96 ± 17 |
| BMI (kg/m<sup>2</sup>) | 34 ± 4.9 | 33 ± 4.7 |
| Hip circumference (cm) | 115 ± 20 | 113 ± 17 |
| Waist circumference (cm) | 116 ± 11 | 114 ± 12 |
| Waist to hip ratio | 1.1 ± 0.7 | 1.0 ± 0.2 |
| Body fat ratio (%) | 42 ± 8 | 39 ± 10 |
| HOMA-index | 2.8 ± 2.1 | 3.4 ± 2.4 |
| Insulin (mU/l) | 10.4 ± 6.4 | 12.1 ± 7.4 |
| Plasma glucose (mg/dl) | 105 ± 20 | 110 ± 20 |
| Hb-A1C (%) | 5.8 ± 0.4 | 5.9 ± 0.7 |
| Hb-A1C IFCC (mmol/mol) | 39.6 ± 4.8 | 41.2 ± 7.4 |
| Triglyceride (mg/dl) | 166 ± 106 | 169 ± 109 |
| Cholesterol (mg/dl) | 220 ± 48 | 222 ± 54 |
| HDL (mg/dl) | 50 ± 11 | 51 ± 10 |
| LDL(mg/dl) | 137 ± 36 | 140 ± 45 |
| LDL/HDL ratio | 2.8 ± 0.7 | 2.8 ± 0.9 |
| CRP (mg/l) | 0.4 ± 0.4 | 0.3 ± 0.3 |
| IL-6 (pg/ml) | 3.1 ± 2.0 | 2.8 ± 2.2 |
| Creatinine (mg/dl) | 0.9 ± 0.2 | 0.9 ± 0.2 |
| eGFR Cockcroft-Gault (ml/min) | 120 ± 39 | 107 ± 32 |

**Fig 1.**
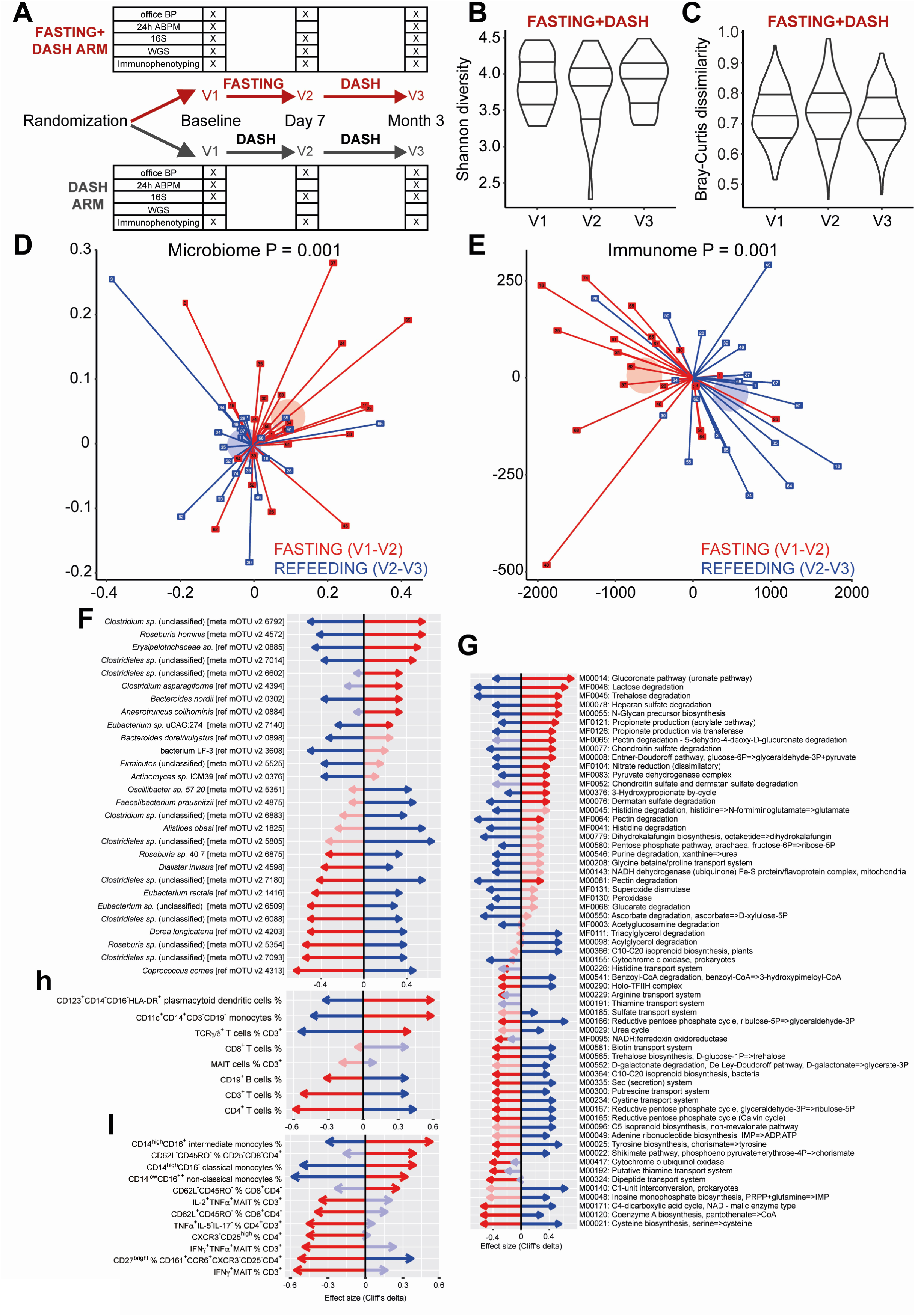
Fasting has pervasive host and microbiome impact. (A) Study design is shown. Subjects are followed from baseline (V1), randomly assigned to begin a modified DASH diet only or to undergo a five-day fast followed by a modified DASH diet. Follow-up is done at one week (V2) and three months (V3). (B) Fasting has no significant (MWU P>0.05) impact on gut microbiome alpha diversity (Shannon diversity from mOTUv2 OTUs) across observation times V1-V3. (C) Fasting has no significant (MWU P>0.05) impact on gut microbiome beta diversity (Bray-Curtis dissimilarity from mOTUv2 OTUs, shown are all between-donor comparisons per time point) across observation times V1-V3. (D) Fasting significantly shifts the gut microbiome towards a characteristic compositional state, while refeeding reverses this change. Unconstrained Principal Coordinates graph with first two dimensions shown. Axes show Bray-Curtis dissimilarities of rarefied mOTUv2 OTUs between samples; each participant in the fasting arm is shown as two lines, one red (fasting change), one blue (refeeding change) connected (centered) at the origin for ease of visualization. Axes show fasting and refeeding deltas after one-week intervention and three-month refeeding. Pseudonym participant ID numbers are shown on the point markers. Transparent circle markers show arithmetic mean position of fasting and recovery deltas respectively. PERMANOVA test P-values reveal significant dissimilarity (P<0.05) between samples from each visit V1-V3 in the original distance space, stratifying by donor. (E) Fasting significantly shifts the host immune cell population towards a characteristic state, while refeeding reverses it. Same as in (D) but using Euclidean distances. (F) Gut microbial taxa significantly enriched/depleted upon fasting/refeeding. Taxa (mOTUv2 OTUs) are shown on the vertical axis, and effect sizes (Cliff’s delta) shown on the horizontal axis. Red arrows represent fasting effects (V2-V1 comparison), blue arrows refeeding effects (V3-V2 comparison). Bold arrows are significant (nested model comparison of a linear model for rarefied abundance of each taxon, comparing a model incorporating patient ID, age, sex and all dosages of relevant medications) to a model additionally incorporating time point, requiring likelihood test Benjamini-Hochberg corrected FDR<0.1 and additionally pairwise post hoc MWU test P<0.05). (G) Gut microbial gene functional modules (KEGG and GMM models analysed together) significantly enriched/depleted upon fasting/refeeding. (H) General immune cell populations significantly enriched/depleted upon fasting/refeeding. (I) Specific immune cell subpopulations. (G-I) Same test as in (F), subset of altered features shown for clarity.

## Results

### Fasting affects the gut microbiome and immunome

As we have previously reported a major influence of common MetS drugs on the microbiota^9^, we accounted for any changes in medication regime or dosage in our statistical tests, alongside controlling for important demographic features such as age and sex. Neither fasting nor refeeding significantly (Mann-Whitney *U* (MWU) P>0.05) altered microbiome species richness/alpha diversity (Shannon, *Figure 1B, Supplementary Figure S1*), or intersample gut taxonomic variability/beta diversity (intersample Bray-Curtis distance, *Figure 1C*; similar results achieved on 16S data (results not shown)). However, there were substantial and significant (PERMANOVA P=0.001) differences in microbial composition within individuals during fasting, reflecting a characteristic intervention-induced shift, which later partially reverted upon a three-month refeeding on a DASH diet (*Figure 1D, Supplementary Table S1 and Figure S2A*). This was echoed by analogous significant (PERMANOVA P=0.001) changes in host immune cell composition during the intervention, revealing a fasting-specific signature, which likewise largely reversed during refeeding (*Figure 1E, Supplementary, Table S1*). DASH without fasting neither affected the microbial composition nor the host immune cell composition (P=0.72 and P=0.38, respectively *Supplementary Figure S2B-C*).

Fasting resulted in a reduction of CD3^+^, CD4^+^ T cells and CD19^+^ B cells, while the frequency of CD8^+^ T cells was unaltered. In contrast, fasting increased the abundance of monocytes (CD14^+^CD11c^+^CD19^-^CD3^-^) and TCRγ/δ^+^ T cells. However, these changes were reversed upon refeeding (*Figure 1H, Supplementary Table S2*). Of note, frequency of CD123^+^CD14^-^CD16^-^ HLA-DR^+^ plasmacytoid dendritic cells also increased upon fasting and was still enriched after refeeding (*Figure 1H, Supplementary Table S2*). When looking closer into monocyte subsets, fasting increased (and refeeding reduced) the frequency of classical CD14^high^CD16^-^, non-classical CD14^low^CD16^++^, and intermediate CD14^high^CD16^+^ monocytes (*Figure 1I, Supplementary Figure S3, Table S2*), which was confirmed by unbiased FlowSOM analyses (*Supplementary Figure S4A-D*). Fasting also affected the relative abundance of differentially activated T cells. Upon fasting, CD8^+^ T cells showed a higher percentage of terminally differentiated cells (T_eff_, CD45RO^-^CD62L^-^) and a lower percentage of the naïve phenotype (T_n_, CD45RO^-^CD62L^+^), while memory T cells were not affected (*Figure 1I, Supplementary Figure S3, Table S1*). A similar pattern was observed in CD4^+^ T_eff_ (*Figure 1I, Supplementary Table S1*). Further, fasting decreased the frequency of pro-inflammatory Th17 (CD27^bright^ CD161^+^CCR6^+^CXCR3^-^CD25^-^CD4^+^), as well as TNFα- and IFNγ-producing Th1 cells (*Figure 1I, Supplementary Table S1*). These changes were partially reverted upon refeeding (*Figure 1I*). Neither fasting nor refeeding changed the overall frequency of CD161^+^Vα7.2^+^ CD3^+^ mucosa-associated invariant cells (MAIT, *Figure 1H, Supplementary Figure S3*). However, frequency of pro-inflammatory MAITs producing TNFα and IFNγ significantly decreased upon fasting and were minimally affected by refeeding (*Figure 1I, Supplementary Table S2*).

Next, we tested all gut microbial taxa and gene functional (KEGG^10^, GMM^11^) modules for abundance shifts during fasting or refeeding, as well as persistent shifts across the three-month study period, controlling for age, sex and any changes in medication (*Figure 1F-H, Supplementary Table S1*). Fasting stimulated shifts in abundance of several core commensals, which was reversed upon refeeding (*Figure 1F, Supplementary Table S1*). Many Clostridial Firmicutes shifted significantly in abundance, with an initial decrease in butyrate producers such as *F. prausnitzii, E. rectale* and *C. comes*, which had also reverted after three months. Interestingly, *C. comes* abundance change was predominantly driven by changes in BMI. Bacteroidaceae showed the opposite pattern and an OTU identified as an *Odoribacter* species likewise bloomed during fasting. At the end of the refeeding period, a persistent depletion could be seen in Enterobacteriaceae, especially *Escherichia coli*. Accompanying the shift in composition are vast changes in microbial metabolic capacity (*Figure 1G, Supplementary Table S1*). Fasting enriches for propionate production capacity, mucin degradation gene modules and for diverse nutrient utilization pathways.

Reanalyzing previously published data, we compared the microbiome signatures of metformin use and MetS to those seen in our novel dataset^9, 12^. For ease of comparability, we proceeded with only human gut specific functional modules (GMM) assessed from shotgun sequencing data available for the fasting arm. Certain fasting- or refeeding-associated functional gene modules from our data were found to overlap with signatures of metformin usage or MetS, but there was little concordance on a taxonomic composition level, in line with previously described higher functional than taxonomic concordance between microbiomes. Of note, when comparing the metformin signal to the MetS signal, it is clear that these two effects are functionally distinct and often oppose one another. Where there were overlapping functional features with results from our novel cohort, the fasting effect appeared to be metformin-like, and during recovery metabolic changes were again reverted (*Supplementary Figure S5*). That is, certain gut metabolic capacities are promoted both by fasting and by metformin treatment.

### Fasting reduces long-term systolic blood pressure and body weight in MetS patients

Assessing the clinical relevance of our intervention, we inspected clinical outcomes in the two study arms. While DASH reduced office SBP after three months (*Figure 2H*), it did not significantly (MWU P=0.27) affect 24h ambulatory SBP, the gold standard of clinical BP measurements (*Figure 2A*)^3^. In contrast, fasting followed by a modified DASH diet led to a sustained reduction both in 24h ambulatory SBP and mean arterial pressure (MAP) (MWU P<0.05, *Figure 2A*). Further, subjects undergoing fasting could significantly (χ^2^ P=0.035) reduce their intake of antihypertensive medication in 43% of cases, compared to only 17% of the cases on DASH alone, while their BP remained under control (*Figure 2B, Supplementary Table S2*). Because the BP response to fasting was heterogenous in our cohort (*Figure 2A-B*), we applied a decision tree model to stratify patients based on their ambulatory BP response, adjusted for antihypertensive medication (*Supplementary Figure S6, Table S3*). The responder group had a median SBP decrease of 8.0 mmHg including a high reduction of medication, while the decrease in the non-responder group was significantly lower (0.3 mmHg; *Figure 2C*). In the DASH only arm, 17 patients were classified as responders with a median SBP decrease of 8.0 mmHg, while the non-responders (n=14) showed no decrease in median SBP (0.5 mmHg, Figure 2C). Fasting followed by a modified DASH diet, unlike a modified DASH diet alone, significantly (drug-adjusted post hoc P<0.05) reduced BMI and body weight even three months post-fasting (*Figure 2D-E*). Although all participants showed a reduction in body weight, this reduction alone could not explain the long-term ambulatory SBP and MAP changes exclusive to the fasting arm (*Figure 2F-G*), nor the microbiome or immunome changes accompanying it (95% of significant findings retain significance when BMI is added as a predictor to the nested models for longitudinal data, see *Supplementary Table S4)*. Only a minority of significant effects (and only two microbiome effects) observed in the fasting-DASH arm could be replicated in the equally powered DASH-only arm (*Figure 2I*).

**Fig 2.**
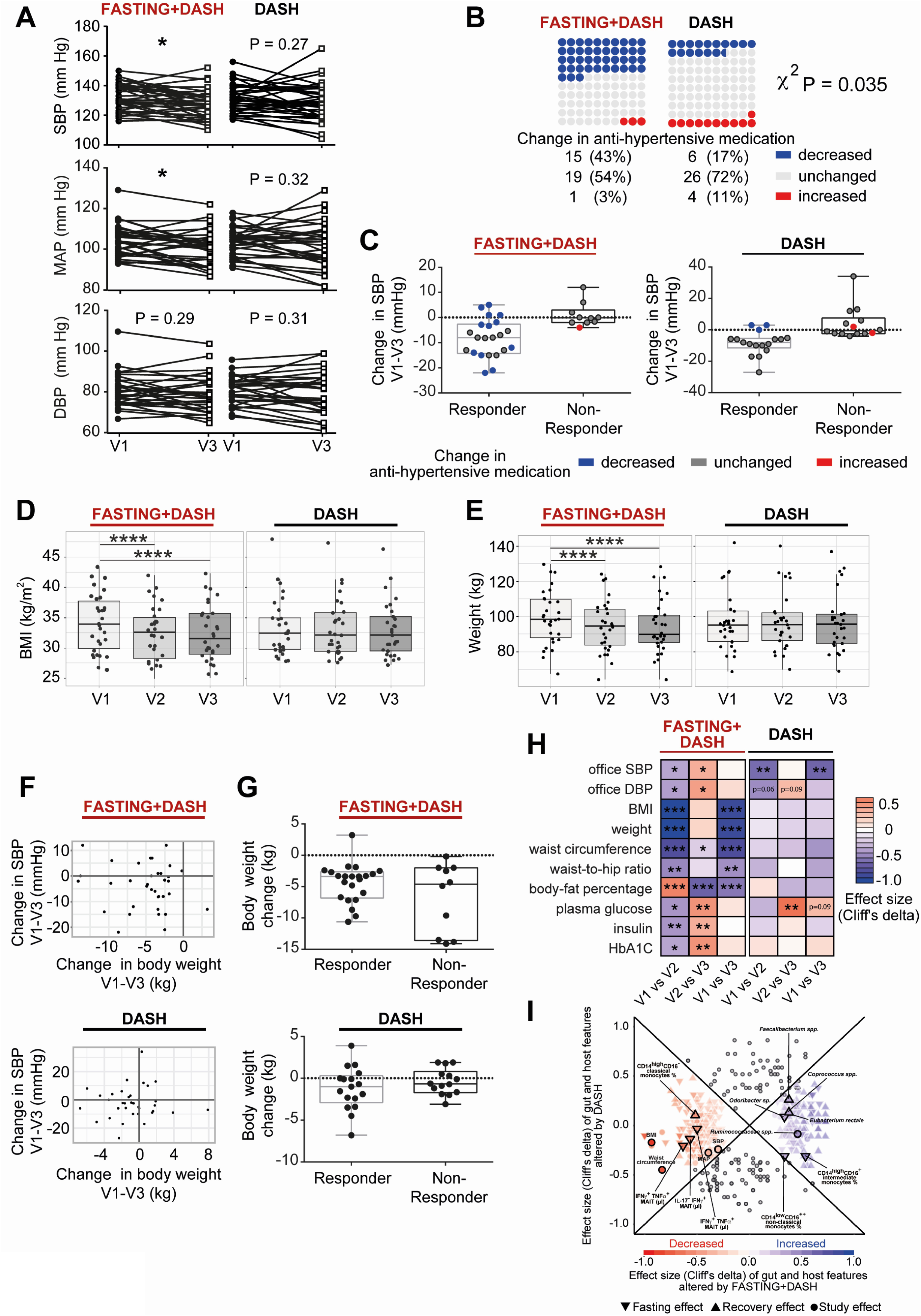
Fasting effects are distinct from those of a modified DASH diet only, and connected to vascular health benefits. (A) Fasting followed by a modified DASH diet, but not a DASH diet alone, significantly (MWU P < 0.05) improves 24h ambulatory SBP and MAP three months post intervention. Lines show individual participant trajectories. (B) MetS subjects beginning a modified DASH diet post fasting significantly reduce their intake of antihypertensive medication by three months post intervention, compared to subjects beginning a DASH diet only. χ^2^ P=0.035. (C) Changes in 24h ambulatory SBP in responders and non-responders including change in antihypertensive medication. (D, E) One week of fasting followed by modified DASH diet, but not DASH diet alone, caused significant (MWU P<0.05) BMI and body weight reduction in MetS patients, persisting three months later. (F) Comparison of changes in 24h ambulatory SBP and body weight, respectively between baseline and followup in both study arms. Each dot represents an individual. (G) Body weight change is not significantly different between responders and non-responders in the fasting arm between baseline and followup. (H) Selected cardiometabolic risk parameters (vertical axis) altered in the fasting arm compared to the DASH arm. Heatmap hues show Cliff’s delta signed effect sizes, with asterisk indicating post hoc univariate significance after compensating for drug dosage changes (see Methods). Horizontal axis shows each time point comparison: change during fasting/week three of DASH, change during refeeding/three months of DASH, and change during the study period as a whole. (I) A majority of host and microbiome effects reported from the DASH+fasting arm do not replicate under DASH only. Comparative effect size plot contrasting features altered significantly only under DASH+fasting (coloured markers, N=275) with features altered significantly also under DASH alone, or with absolute effect size greater in DASH alone (gray markers, N=133). For the former category, colour hue shows direction of effect, colour intensity scope of effect, and marker shape which time point comparison is shown. Vertical axis shows effect size in DASH only, horizontal effect size in DASH+fasting. Selected features are named for reference.

### BP responder-specific changes in the gut microbiome and immunome

Because the BP responsiveness was heterogenous in the fasting-DASH arm (*Figure 2A-C*), we hypothesized that unique characteristics involving the immunome or microbiome of these patients may contribute to their BP response. We compared the impact of fasting and refeeding in the complete fasting arm, in the BP-responders of the fasting arm, and in the DASH-only arm (*Figure 3A-B, Supplementary Table S1, 5, 6*). Even at reduced statistical power, we were able to capture changes in abundance of many gut microbial taxa that were uniquely characteristic of successful fasting treatment, most of which displayed the fasting-refeeding reversal dynamic described above (*Figure 3A, Supplementary Table S5, 6*). Responders were especially characterized by an initial and sustained enrichment of an unclassified *Odoribacter* species with concomitant *Actinomyces* depletion. Responders experienced an initial depletion, and later a strong enrichment, of the butyrate producer *F. prausnitzii*. Accordingly, in BP-responders, fasting resulted in an initial bloom and later depletion of *Bacteroides* (*Figure 3A*).

**Fig 3.**
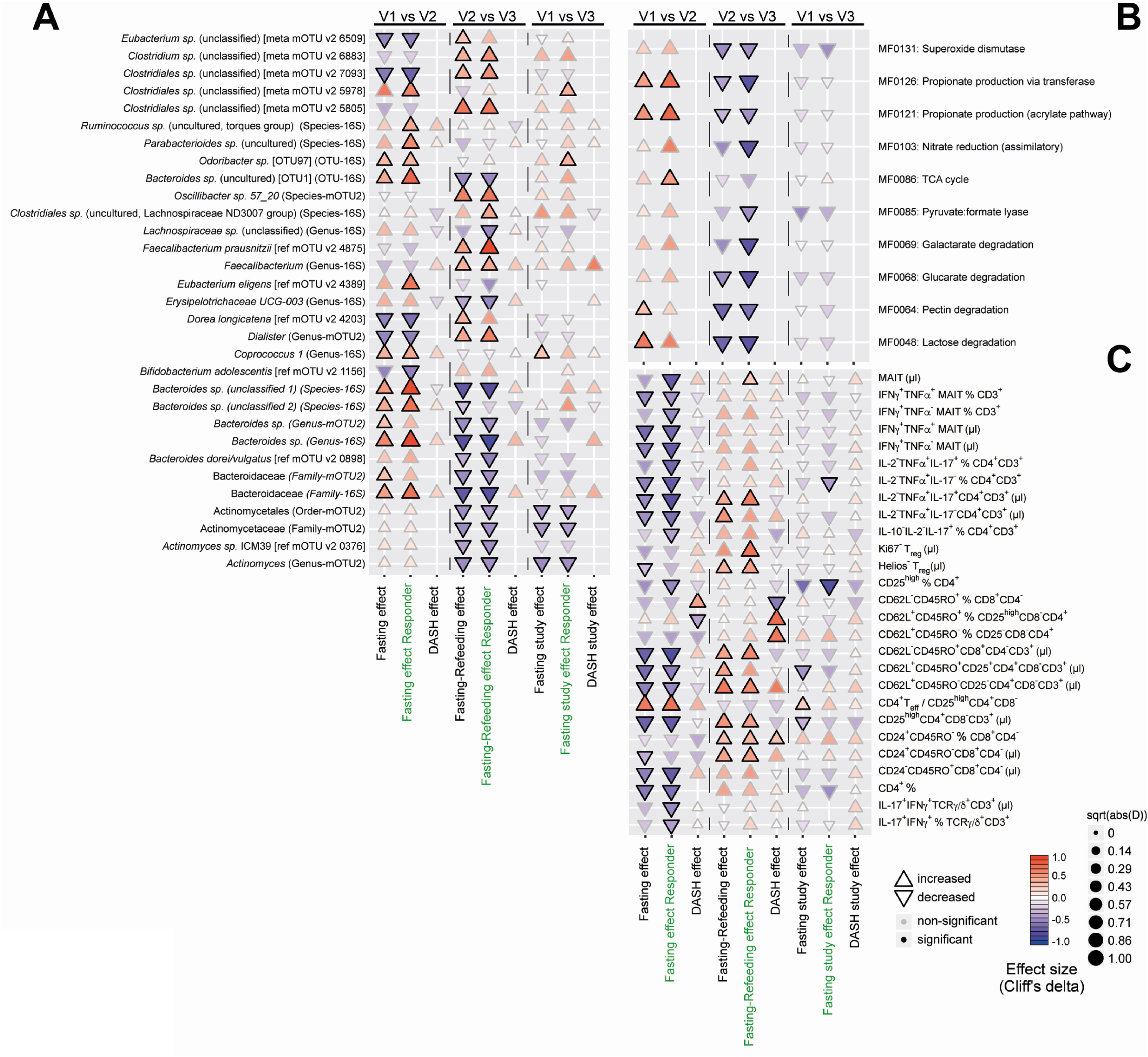
Subjects responding favorably to fasting exhibit stronger changes in commensal abundance under intervention. (A) Cuneiform plot shows subset of bacterial taxa, at different taxonomic levels and measured either using 16S sequencing or shotgun sequencing, altered significantly (drug-adjusted post hoc FDR<0.05) in abundance tested in intervention responders only (vertical axis). Signed effect size are shown through marker direction and color, hue and size represent absolute effect size. Solid borders indicate significance. Markers not shown could not be tested in the DASH arm as shotgun data was unavailable, or showed no difference in rank-transformed values (Cliff’s delta=0). Horizontal axis separates tests for fasting (comparison of baseline to after one week), recovery (comparison of after one week to three months), and study effect (comparison of baseline to three-month follow-up). DASH results are from the DASH arm only, responders are tests using only the responders (as per decision tree) in the fasting arm. (B) Same view as (A) but with regards to gut functional modules (selected subset shown for clarity). (C) Same view as (A) but with regards to immune cell subpopulations (selected subset of GMMs shown for clarity). T_reg_: FoxP3^+^ cells, MAIT: Vα7.2^+^CD161^+^CD4^-^ CD3^+^.

Virtually no overlap with effects seen in the equally powered DASH arm were found, indicating that fasting is needed on top of a BP-reducing diet for these changes to occur (*Supplementary Figure S7*).

In profiling the microbial metabolic potential in BP-responders, we focused on gene modules curated for relevance to metabolism in the human gut (GMM)^11^. On a functional level, responder-characteristic changes resemble those in the fasting arm at large, but with even more pronounced relative enrichment for propionate production (MF0126, MF0121) modules (*Figure 3B*). Some modules were significantly altered in abundance only in this stratified subgroup, indicating these changes strongly characterize responders compared to non-responders (*Supplementary Table S5*). For example, pyruvate:formate lyase (MF0085) is depleted during recovery only in responders.

Changes to the immunome of responders are similar to those seen in the unstratified fasting group and differ from those in the DASH arm (*Figure 3C, Supplementary Table S1, 5, 6*). In the fasting arm, several immune features related to pathogen-sensing and mucosal immunity (e.g. MAIT cells, IL-17^+^-producing Th and γδT cells) changed significantly in abundance only when tested in the responder group, indicating relevant differences between responders and non-responders. Upon fasting, frequency of both pro- and anti-inflammatory adaptive immune cells showed a stronger decrease in responders, indicating a stronger anti-inflammatory effect of fasting in responders.

### Network analysis of microbial, immune, and clinical features

We next aimed to study interacting microbiome-immune features through network analysis, seeking to explain the beneficial role of fasting on BP. We assessed all triplets of pairwise interactions between host clinical phenotypes, immune cell populations, and microbiome taxa or functional profiles, respectively, using modified Spearman correlations (requiring FDR<0.1 in each comparison of two data spaces, and P<0.05 in a post hoc test accounting for the presence of the same subjects at all three time points (see Methods, *Supplementary Table S7*). *Figure 4A* shows a chord diagram constructed from these data, where the colored outer rings are lined with components from one of our three tested system spaces during fasting, refeeding and over the full duration of the study, and where the color of the connectors between factors indicate a positive or negative association (Spearman’s rho). We identified a cluster of circulating cytokine-producing MAIT cells (absolute number and fraction of CD3^+^ T cells), which positively correlated with 24h ambulatory SBP (*Figure 4A, E, Supplementary S8*) and MAP, but not with 24h diastolic BP (*Figure 4A, Supplementary Table S7*). In our patients, a number of IL-2^+^TNFα^+^ producing-MAIT cells positively correlated with the abundance of a species of *Firmicutes* (unclassified) [meta_mOTU v2 6331] and negatively with Bifidobacteriales (Order-16S), while IFNγ^+^ producing-MAIT cells positively correlated with abundance of *Barnesiella intestinihominis* [ref_mOTU v2 4880], *Clostridiales sp*. (unclassified) [meta_mOTU v2 6787] and *Bacteroides eggerthii* [ref_mOTU v2 1410] (*Figure 4A, D, Supplementary Table S9*). Abundance of various microbes including *Coprococcus sp*. were negatively associated with IFNγ^+^ producing-MAIT cells (*Figure 4A, D, Supplementary Table S9*). Interestingly, fasting depleted various cytokine producing-MAIT cells with the most pronounced long-lasting decrease seen for IL-2^+^TNFα^+^ producing-MAIT in BP-responders (*Figure 4A-C, Supplementary Table S9*).

**Fig 4.**
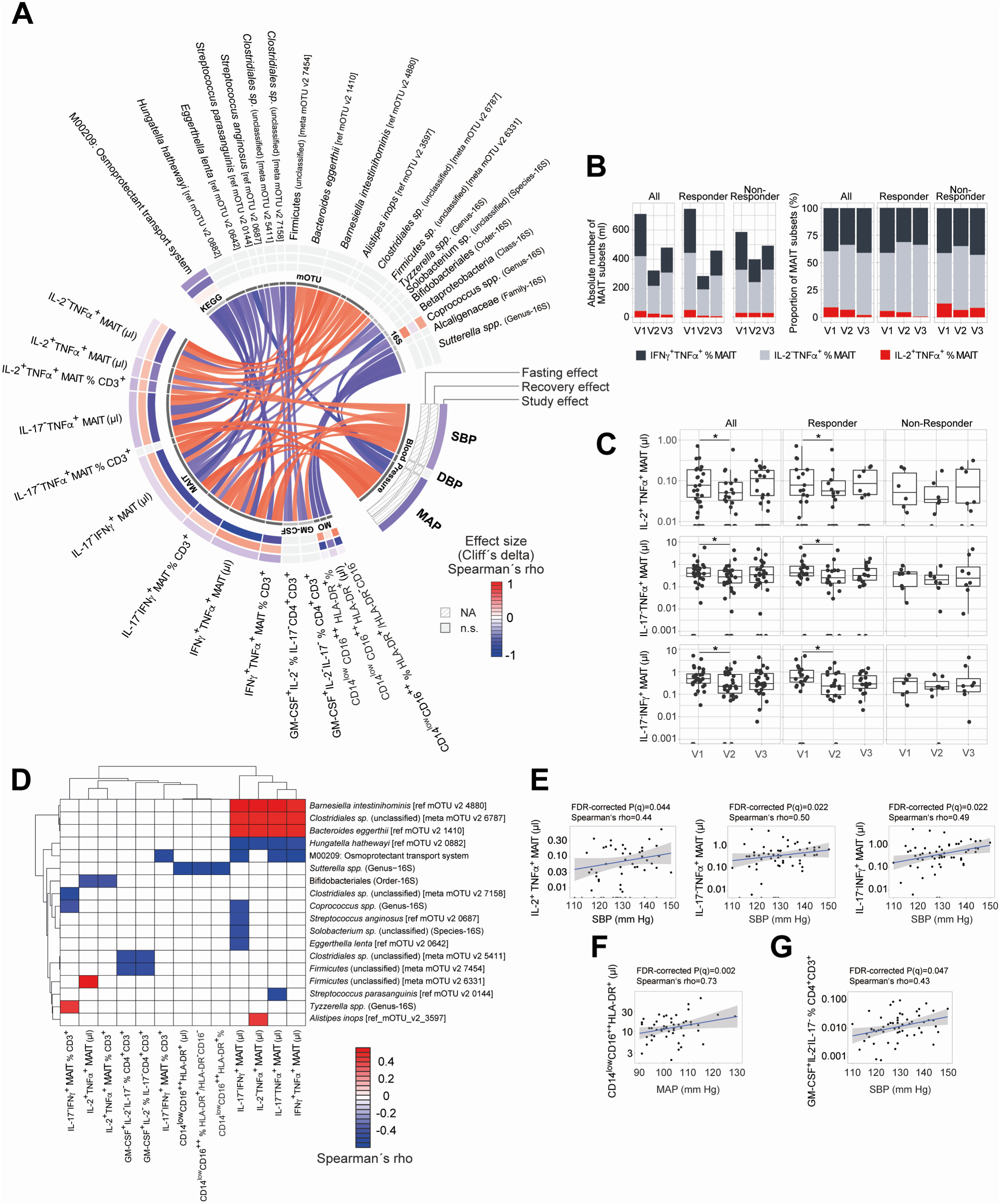
Blood pressure-microbe-immune association. (A) Chord diagram visualises the interrelation between BP (24h ambulatory systolic, mean or diastolic BP) and fasting-impacted microbiome functional or taxonomic features, and immune cell subsets. Features are shown that form triplets of immune, microbial and phenotype variables where at least two of three correlations are significant (Spearman FDR < 0.05, post hoc nested model test accounting for same-donor samples < 0.05) in the fasting arm of our novel cohort, and where in addition one or more feature significantly (drug-adjusted post hoc FDR < 0.05) are affected by the intervention. Colour of the connectors indicates positive or negative association (Spearman’s rho), colour of the cells within the tracks indicates changes upon fasting, refeeding and study effect (Cliff’s delta, white if not significant), respectively. (B) Cumulative absolute number and relative abundance of circulating IFNγ^+^TNFα^+^, IL-2^-^TNFα^+^ and IL-2^+^TNFα^+^ mucosa-associated invariant T cells (MAIT) cells within the fasting arm subdivided by BP-responsiveness (median, n=30 for all, n=20 for responders, n=8 for non-responders, respectively). (C) Absolute number of circulating IL-2^+^TNFα^+^ (All: P = 0.019, Responder: P = 0.024), TNFα^+^ (All: P = 0.006; Responder: P = 0.022) and IFNγ^+^ (All: P = 0.001; Responder: =0.007) MAIT cells within the fasting arm subdivided by BP-responsiveness (n-number as in (B); MWU test after Benjamini-Hochberg correction). (D) Hierarchical clustering of microbiome features-associated immune features. Colour indicates Spearman’s rho. (E) Correlations of circulating IL-2^+^TNFα^+^, TNFα^+^ and IFNγ^+^ MAIT cells and 24h ambulatory SBP (*FDR-corrected P=0.044, 0.022 and 0.022, respectively). (F) Correlations of circulating non-classical CD14^low^CD16^++^HLA-DR^+^ monocytes and 24h ambulatory MAP in responder (*FDR-corrected P=0.002). (G) Correlations of circulating GM-CSF^+^IL-2^-^IL-17^-^ of % CD3^+^ and 24h ambulatory SBP (*FDR-corrected P=0.047). n-number for (C-G) as in (B). MAIT: Vα7.2^+^CD161^+^CD4^-^CD3^+^.

The association between MAIT cells and BMI is still a matter of debate^13^. We found in our study that the abundance of MAIT cells did not correlate with either BMI, weight, waist circumference, waist-hip ratio or body fat percentage (*Supplementary* Figure S8, *Table S7*). However, we found that these weight-related parameters did correlate with the abundance of a subset of circulating Treg-like cells (CD62L^-^CD45RO^-^CD25^+^CD4^+^), a cell type previously linked to morbid obesity in human subjects^14^.

A recent publication showed non-classical monocyte enrichment in hypertensive patients^15^. Interestingly in our study, circulating non-classical monocytes were enriched upon fasting and then depleted again upon refeeding to remain below baseline levels three months after fasting (*Figure 1H, 4A, Supplementary Table S1)*. Network analysis revealed an association between non-classical monocytes, MAP and gut abundance of Betaproteobacteria. *Sutterella spp*., belonging to the class Betaproteobacteria, showed an inverse correlation with non-classical monocytes, while *Clostridiales* [meta_mOTU_v2_6233] was positively associated with non-classical monocytes and MAP (*Figure 4A, D, F, Supplementary Table S7*). We found that the frequency of circulating GM-CSF-producing CD4^+^ T cells was negatively correlated with unclassified *Firmicutes* [meta_mOTU v2 7454] and *Clostridiales sp*. (unclassified) [meta_mOTU v2 5411], and positively with SBP (*Figure 4A, D, G, Supplementary Table S7*).

### Baseline indicators predicting efficacy of fasting on blood pressure

As previously stated, a large proportion of fasting patients responded with a substantial drop in BP, allowing them to reduce their use of anti-hypertensive medication while BP remained controlled. As not all patients experienced this beneficial effect, we sought to understand whether the factors underlying successful fasting intervention in the BP-responders could be predicted at baseline. Responder and non-responder subgroups differ considerably in immunome and microbiome features, not only post-fasting and at three-month follow-up, but also at baseline, suggesting a favorable clinical response may be predictable in single patients (*Supplementary Figure S9A-C*). Regarding responder-specific features, we identified a small number of features as both characteristic of responders at baseline and during the intervention (*Figure 5A*). Microbiomes of BP responders were depleted pre-intervention for two poorly characterized taxa (*Bacteroides sp*. and *Parabacteroides sp*.) and for propionate biosynthesis (*Figure 5A*). Fasting strongly elevated the abundance of both taxa and enriched these propionate production modules, indicating that responders suffer a treatable deficit. By three months post intervention, propionate modules and *Bacteroides sp*. are almost back at baseline while BP (relative to medication dosage) remains improved, suggesting that their transient elevation during refeeding may have stabilized a less hypertensive state through downstream mechanisms. In addition, the difference in abundance of *Parabacteroides sp*. in responders and non-responders had equalized by time of follow-up.

**Fig 5.**
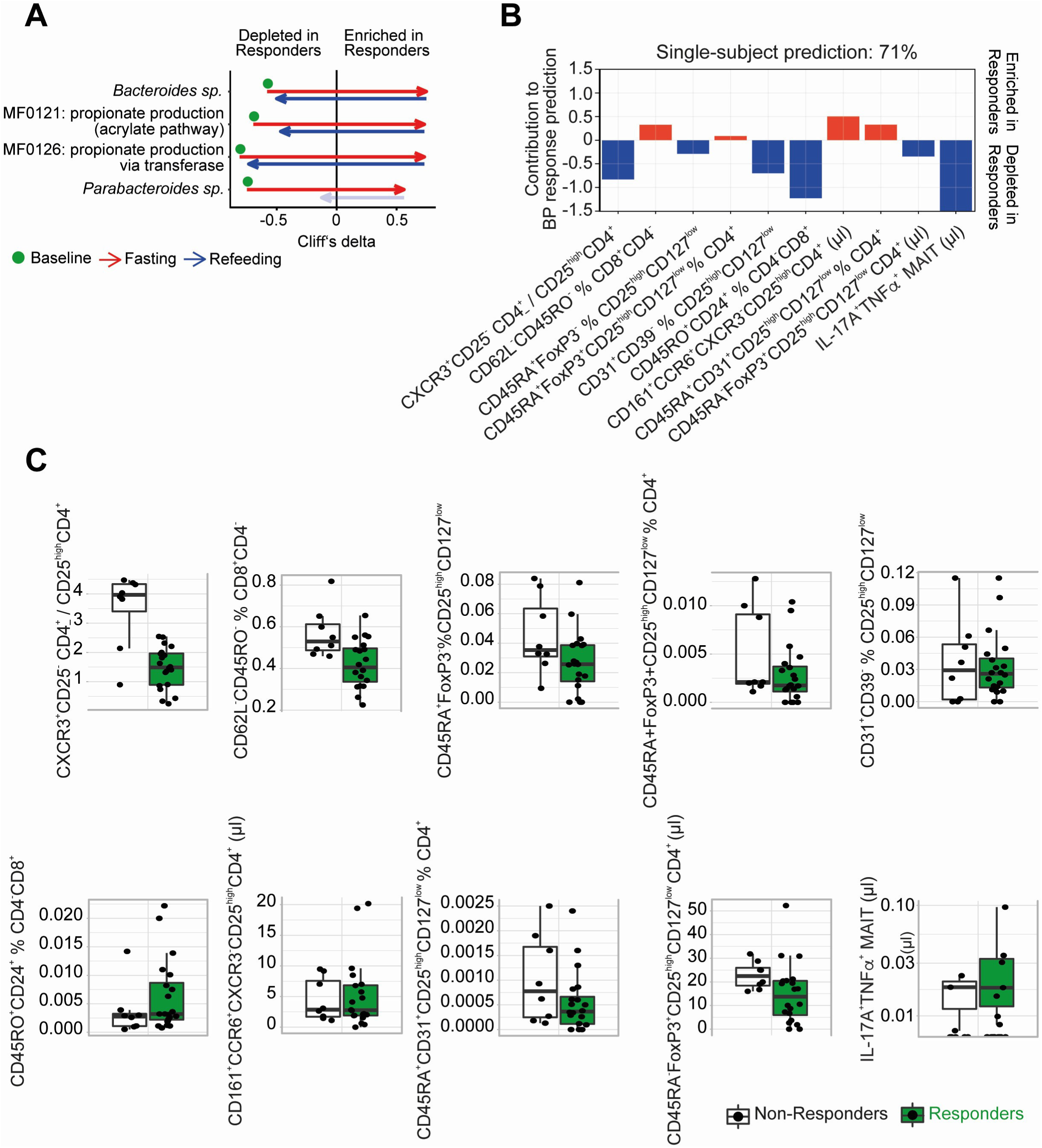
Long-lasting BP-responders and non-responders differ in microbiome and immunome composition. (A) Features altered at baseline in responders vs. non-responders and altered during intervention in responders. Effect size (Cliff’s delta) is shown comparing responders and non-responders. Transparency indicates non-significant differences (drug-adjusted post hoc FDR≥0.05). (B) Prediction model weights for blood pressure response using the immunome dataset at baseline. The top ten immunome features were used to build a multivariate logistic-regression algorithm. Single-subject prediction was quantified using a leave-one-out cross-validation procedure. The bar plots represent the regression in a model with binary output (responder yes=1 vs no=0) for every feature. (C) Quantification of the immunome features at baseline used in the prediction model to predict BP response in the future, split into responders and non-responders. MAIT: Vα7.2^+^CD161^+^CD4^-^CD3^+^.

As mentioned above, immunome composition also differed already at baseline in responders and non-responders. To further elucidate this phenomenon, we applied machine learning algorithms and empirically show that we can make effective predictions from the immunome data. From 494 total immune variables, step-wise forward regression identified the top ten discriminators of responders from non-responders at baseline. A prediction accuracy of 71% (sensitivity 75%, specificity 70%, and F1 score 77%) was achieved using a leave-subject-out cross-validation for whether or not a future patient would respond favorably to fasting with regards to BP (*Figure 5B*). Within this multivariate analysis, the driving immune features of this classifier highlighted a lower CXCR3^+^CD25^−^CD4^+^/CD25^high^CD4^+^ (most likely Th1/Treg ratio), alongside lower abundances of CD24^+^ memory CD8^+^ T cells and IL-17^+^TNFα^+^MAIT cells in responders relative to non-responders (*Figure 5C, Supplementary Figure S9E*). Regarding the top ten features derived as indicative for successful patient classification, responders seem to have less of a pro-inflammatory immune signature at baseline (*Figure 5C*). Notably, we could increase the prediction performance of the classifier up to 78% by using changes of immune cell abundances between baseline and three-month follow-up visit as a basis for prediction of BP response at the single-patient level (Supplementary *Figure S9D, F*). In contrast, for subjects on a DASH diet only, corresponding classifiers were unable to predict BP response above chance level.

## Discussion

In summary, we have carried out the first high-resolution multi-omics characterization of fasting in patients with metabolic syndrome. The major novel finding is that periodic fasting followed by 3 months of modified DASH diet induces concerted and distinct microbiome and immunome changes that are specific to fasting itself leading to a sustained BP benefit (Fig. 2I), since it cannot be reduced by DASH diet alone. Fasting followed by modified DASH led to a significant long-term reduction in body weight. However, neither the change in blood pressure nor global changes to the microbial composition or immunome appear mediated by this BMI decrease (95% of findings retain significance when deconfounding for BMI change, see *Table S4*, and body weight reduction is not more pervasive in treatment responders than nonresponders, *Figure 2G*). This suggests that further sustained weight loss beyond what is observed here may synergize with the fasting benefits to reduce hypertension even further. Fasting induced a profound change in circulating immune populations; e.g. depleted Th1 cells and permanently enriched dendritic cells, which both have been shown previously to play a role in the pathogenesis of experimental hypertension^16, 17^. Further, we discovered significant correlations between circulating MAIT cells and 24h ambulatory BP and MAP.

A growing body of evidence suggests that the abundance of certain microbes is associated with cardiovascular health. Previous reports on hypertensive patients show taxonomic and functional gut microbiome shifts^6, 7^. For example, *Firmicutes* have been shown to be more abundant in healthy controls compared to pre-hypertensive and hypertensive patients^7^. Upon fasting, several Clostridial Firmicutes shifted significantly in abundance, with an initial decrease in butyrate producers such as *F. prausnitzii, E. rectale* and *C. comes*, which were also reverted after three months upon refeeding; with the latter taxon one of few likely to be indirect effects of the observed weight reduction (*Table S4*). Bacteroidaceae showed the opposite pattern, and an unclassified *Odoribacter* species likewise bloomed during fasting. Of note, *Odoribacter sp*. has been negatively associated with both SBP^18^ and vascular stiffness^19^ in obese women. *Escherichia coli* was shown to be associated with endothelial dysfunction and MetS. In our study, fasting permanently depleted *Escherichia coli*, suggesting that fasting could promote cardiovascular health through the gut microbiome. Further, functional microbial metabolism in fasting patients at baseline share some similarities to the previously profiled hypertensive microbiome^7^. In the fasting arm, the functional shift during refeeding enriches for functional modules also enriched in non-hypertensive controls, i.e. for potentially BP-protective factors. Clinical studies represent a highly heterogeneous situation with multifactorial disease features and strongly variable microbial and lived environments. To account for this heterogeneity, we compared the data from our longitudinal study (post-fasting and 3-month) to the respective baseline values of the study subjects. This intraindividual analysis allowed us to identify BP responder-specific changes despite even reduced power in such a sub-stratified analysis. Notably, despite the non-homogenous starting conditions in our MetS patients, our five-day fasting followed by a DASH intervention improved BMI in all participants and ambulatory SBP in two thirds, even three months post-intervention. The responder-specific microbiome changes in our fasting arm post-intervention (enrichment of an *Odoribacter* species, *Bacteroides* and *Firmicutes*, depletion of Actinomyces and *F. prausnitzii*) are likely beneficial to the host. A recent study profiling the hypertensive microbiome showed that during disease, patients experienced an enrichment of Actinomyces, and a depletion of *F. prausnitzii, Bacteroides* and Firmicutes^7^. In addition, some functional gut specific gene modules in our dataset were enriched only in BP-responders, for example the pyruvate:formate lyase module, MF0085. Our data are congruent with a study showing enrichment of the same enzyme in symptomatic atherosclerosis patients compared to healthy controls^20^.

The fact must also be addressed as to how, despite the fact that many of the immunome and microbiome shifts post-fasting are transient, there exists a sustained improvement of cardiovascular health in our patients. We hypothesize that during a specific time window during early refeeding, certain fasting-depleted taxa (including core butyrate producers) and associated immune cells may be enriched beyond both baseline or follow-up at 3 month, leading to a temporary state of high butyrate availability, especially on a DASH diet. Therefore, we speculate that anti-inflammatory effects of this SCFA during regrowth, as well as similar effects of propionate during fasting itself, may be important mechanisms behind the sustained improvement in blood pressure that we observe. While we cannot directly test it in the present cohort, it is a scenario consistent both with expectations from the literature and with our observations of a consistent depletion-then-regrowth pattern. Thus, future work will involve studying a fasting/refeeding cohort also at intermediate time intervals. It is likewise a limitation that we cannot infer direction or causality of concomitant immunome-microbiome changes from the present cohort study alone. Accordingly, our next steps will also involve direct manipulation of microbiota, as well as direct infusion of their potentially immunomodulating metabolites, in animal models for hypertension, similar to some of our recent proof-of-principle studies^21, 22^.

Stratification of the cohort to BP-responsiveness showed that also immune changes present in the fasting arm are more pronounced in responders than in non-responders, and are fundamentally different from the changes observed in the DASH-only arm. The DASH-only arm was associated with the decrease of CD8^+^ Tem cells, previously reported to play a role in hypertension^17, 23^. Responders and non-responders not only reacted differentially to fasting, but also differed at baseline, including by depletion of *Bacteroides sp*., *Parabacteroides sp*., and propionate synthesis genes pre-intervention. These features were then normalized during fasting. Notably, recent experimental work suggested an anti-hypertensive effect of propionate treatment in mice.^22^ Furthermore, *Parabacteroides* are more commonly found in the microbiome of hypertensive subjects than in healthy controls^6^. Our findings indicate responders and non-responders to our intervention differ with regards to several gut microbiome features relevant to hypertension, with fasting-induced normalization of these differences seen during a successful fasting intervention.

Through network analysis of the immunome, microbiome and clinical data, we identified significant correlations between circulating MAIT cells and 24h ambulatory BP and MAP. MAIT cells represent up to 10% of peripheral blood T cells, but in contrast to other classical T cells^17^, have not yet been linked to the regulation of BP. They differ in many aspects from conventional T cells by expressing a semi-invariant TCR α-chain Vα7.2-Jα33. MAITs can produce various cytokines mimicking an effector/memory-like phenotype and yet they behave rather like innate cells. During aging^18^ and CMD^13, 24^, absolute circulating MAIT number and frequencies decrease, while certain subsets of cytokine producing and adipose tissue MAITs were found to be enriched in obese type 2 diabetic patients^13^. In contrast to a previous report showing significant enrichment of IL-17-producing MAIT cells (after re-stimulation) in severely obese patients (average BMI=54^13^), our cohort (average BMI=34) had very low IL-17 producing MAITs, and they were only quantifiable in a small group of patients (data not shown).

Using machine learning, we were able to utilize deep immunophenotyping data to predict at baseline which subjects were likely to decrease their BP during fasting despite the small number of subjects. In addition, accuracy of the prediction was enhanced further taking the dynamics of immune populations along the course of the study into account. No corresponding prediction of favorable response to a DASH-only intervention was possible. Thus, we demonstrate the practical utility of a machine learning analysis pipeline for predicting BP benefit of fasting in MetS patients with hypertension. Personalized nutrition, informed by -omics measurements, is therefore a possible translational benefit of our study.

## Limitations

It is important to recognize that our study represents patients with hypertension and MetS solely from a Caucasian-European background. This selection criterion introduces a selection bias in our study design. Additional research is necessary to elucidate whether the results presented here could be generalizable to a more heterogenous patient population. Further, our recruitment procedure could already have introduced a selection bias toward patients who were interested in fasting/dietary studies and therefore are sensitive about their cardiometabolic health. Since the participants were especially interested in the fasting procedure, the allocated DASH participants were offered after successful completion of the study a cost-free fasting cycle. However, we cannot exclude that this led to an increased long-term motivation compared to the participants, who started with the fasting protocol. In addition, the relatively low patient number could be regarded as a limitation. Although our present study is large enough to allow inference of significance for most strong contributors to the observed effect, results are likely not complete, and follow-up in additional and larger studies will be needed for a comprehensive view of subtle fasting-associated host and microbiome features. Our study design did not allow to blind participants regarding their intervention. To maximally reduce the bias, the scientific staff were blinded during the process of processing and analyses of collected samples and measurements. Further, the present study cannot infer how frequently fasting cycles should be repeated to control BP in at-risk patients, nor whether it is as effective without a concomitant DASH intervention. Future clinical studies are needed to test these hypotheses.

## Conclusions

Our data demonstrates that fasting induces changes to gut microbiome and immune homeostasis with a sustained beneficial effect on body weight and BP. The favorable impact of fasting shown here highlights this intervention as a promising non-pharmacological intervention for the treatment of MetS.

## Data Availability

The data that support the findings of this study are available from the corresponding author upon reasonable request.

## Acknowledgments

We thank Juliane Anders, Jana Czychi and Gabriele N’Diaye for the outstanding technical assistance, Falk Hildebrand, Luis Pedro Coelho and Renato Alves for assistance with metagenomic software adaptation.

